# Hidden fraction of Polish population immune to SARS-CoV-2 in May 2021

**DOI:** 10.1101/2021.06.22.21258711

**Authors:** Wiktoria Budziar, Katarzyna Gembara, Aleksander Szymczak, Natalia Jędruchniewicz, Krzysztof Baniecki, Aleksandra Pikies, Artur Nahorecki, Agnieszka Hoffmann, Amelia Kardaś, Tomasz Klimek, Zuzanna Kaźmierczak, Wojciech Witkiewicz, Kamil Barczyk, Krystyna Dąbrowska

## Abstract

Population immunity (herd immunity) to SARS-CoV-2 derives from two well-defined and controlled sources: vaccinations or diagnosed and registered cases of the disease. It may however also result from asymptomatic, oligosymptomatic, or even full-blown but undiagnosed and unregistered cases from which patients recovered at home. Here we present a population screening for SARS-CoV-2 specific IgG and IgA antibodies in Polish citizens (N=501) who had never been positively diagnosed with or vaccinated against SARS-CoV-2. Serum samples were collected in Wrocław (Lower Silesia) on 15th and 22nd May 2021. Sera from hospitalized COVID-19 patients (N=22) or from vaccinated citizens (N=14) served as positive controls.

Sera were tested with Microblot-Array COVID-19 IgG and IgA (quantitative) that contain specific SARS-CoV-2 antigens: NCP, RBD, Spike S2, E, ACE2, PLPro protein, and antigens for exclusion cross-reactivity with other coronaviruses: MERS-CoV, SARS-CoV, HCoV 229E Np, HCoV NL63 Np.

Within the investigated population of healthy individuals who had never been positively diagnosed with or vaccinated against SARS-CoV-2, we found that 35.5% (178 out of 501) were positive for SARS-CoV-2-specific IgG and 52.3% (262 out of 501) were positive for SARS-CoV-2-specific IgA; 21.2% of the investigated population developed virus-specific IgG or IgA while being asymptomatic. Anti-RBD IgG, which represents virus-neutralizing potential, was found in 25.6% of individuals (128 out of 501).

These patients, though positive for anti-SARS-CoV-2 antibodies, cannot be identified in the public health system as convalescents due to undiagnosed infections, and they are considered unaffected by SARS-CoV-2. Their contribution to population immunity against COVID-19 should however be considered in predictions and modeling of the COVID-19 pandemic. Of note, the majority of the investigated population still lacked anti-RBD IgG protection (74.4%); thus vaccination against COVID-19 is still of the most importance for controlling the pandemic.

## Introduction

COVID-19 is an acute respiratory disease caused by the novel coronavirus SARS-CoV-2, also known as 2019-nCoV. The World Health Organization (WHO) characterized COVID-19 as a pandemic on March 11^th^ 2020. Since then, the COVID-19 pandemic has affected most aspects of life globally, including drastic lockdowns to reduce the death toll [1], [2], [3]. Extraordinary efforts have been focused on developing drugs and vaccines that could help the situation. A universal anti-COVID-19 drug has still not been developed, and the major strategy for controlling the pandemic is population immunity, with the key role of SARS-CoV-2 targeting vaccines [4].

Population immunity to SARS-CoV-2 derives from two well-defined and controlled sources: vaccinations or diagnosed and registered cases of the disease. It may however also result from asymptomatic, oligosymptomatic, or even full-blown but undiagnosed and unregistered cases from which patients recovered at home. COVID-19 symptoms that are typical to many other flu-like and cold infections make it impossible to identify this disease without specific diagnostics. For this reason there is an unknown fraction of society that has already achieved immunity to COVID-19 but its extent is unknown. Attempts to estimate this fraction in each society is difficult, and it remains a subject of speculation, sometime extreme [5], [6].

Here we present the results of population screening for SARS-CoV-2 specific antibodies in Polish citizens (N=501) who had never been positively diagnosed with or vaccinated against SARS-CoV-2. Blood samples were collected in Wrocław (Lower Silesia) on 15th and 22nd May 2021, which was shortly after the second main wave of the disease in Poland that was observed approximately between 15th February and 30th April 2021, reaching more than 35 000 diagnosed cases per day at its peak (out of almost 38 million citizens).

Overall statistics of COVID-19 for the whole country at 15th May were as follows:

- Total number of all infected with SARS-CoV-2 during the pandemic: 7.5% of population (2 851 911/71 609 total infected/died).
- Vaccinated with at least 1 dose: 30.3% of population, which includes full vaccination: 11.9% of population

Overall statistics of COVOD-19 for Lower Silesia at 15^th^ May were as follows:

- Total number of all infected with SARS-CoV-2 during the pandemic: 7.3% of population (210 353/4634 total infected/died).
- Vaccinated with at least 1 dose: 31.9% of population, which includes full vaccination: 11.8% of population

## Material and methods

### Blood samples

Blood samples were collected in Wrocław (Lower Silesia) on 15th and 22nd May 2021, from healthy individuals (over 17 years old, N=501), who had not been vaccinated against COVID-19 with any kind of vaccine, nor had they ever been diagnosed positively for SARS-CoV-2 infection. The positive control was the group of patients hospitalized due to severe COVID-19 (non-vaccinated) 10-30 days after estimated start of the infection (N=22) and individuals vaccinated against SARS-CoV-2 at least 2 months after the second dose (never diagnosed positively for SARS-CoV-2 infection) (N=14). Serum was collected into standard clotting tubes (BD SST II Advance), left for 1 hour at room temperature (RT) to clot and separated from the clot by centrifugation (15 min, 2000 g, RT), then stored at -20°C for further use. All patients were interviewed for possible flu-like and cold-type infections within the period of the COVID-19 pandemic. All reads and data are fully available for further analyses in the supplementary material (Table S1).

### Bioethics statements

The experiments were approved by the local Commission of Bioethics of the Regional Specialist Hospital in Wrocław (approval number: KB/02/2020, policy No. COR193657).

### Serological diagnostic tests

Microblot-Array COVID-19 IgG and Microblot-Array COVID-19 IgA (TestLine Clinical Diagnostics s.r.o); cat no. CoVGMA96, LOT 0100060496 and cat. no. CoVAMA96, LOT 0100066601); the arrays contain a combination of selected parts of the specific antigens of SARS-CoV-2: NCP, RBD, Spike S2, E, ACE2, PLPro protein, and antigens for exclusion cross-reactivity with other endemic coronaviruses: MERS-CoV, SARS-CoV, HCoV 229E Np, HCoV NL63 Np.

### Statistics

Data were analyzed by ANOVA and the Kruskal–Wallis test with the Statistica 8.0 software package (www.statsoft.pl).

## Results and Discussion

Within the investigated population of healthy citizens who had never been positively diagnosed with or vaccinated against SARS-CoV-2, we found that 35.5% (178 out of 501) were positive for SARS-CoV-2-specific IgG and as many as 52.3% (262 out of 501) were positive for SARS-CoV-2-specific IgA (Table 1). These positive patients (with the exception of 2 individuals) did not demonstrate any cross-reactivity to other coronaviruses; thus their immune response to SARS-CoV-2 is specific and it apparently results from immunization with SARS-CoV-2. Taking the high rate of SARS-CoV-2-specific IgA, more than a half of the investigated population has had a contact (or infection) with the virus. Interestingly, 106 patients positive for SARS-CoV-2-specific IgG or IgA declared no flu-like infection or cold within the time of the COVID-19 pandemic; thus, 21.2% of the investigated population developed virus-specific antibodies while being asymptomatic (Table S1).

**Table 1.**
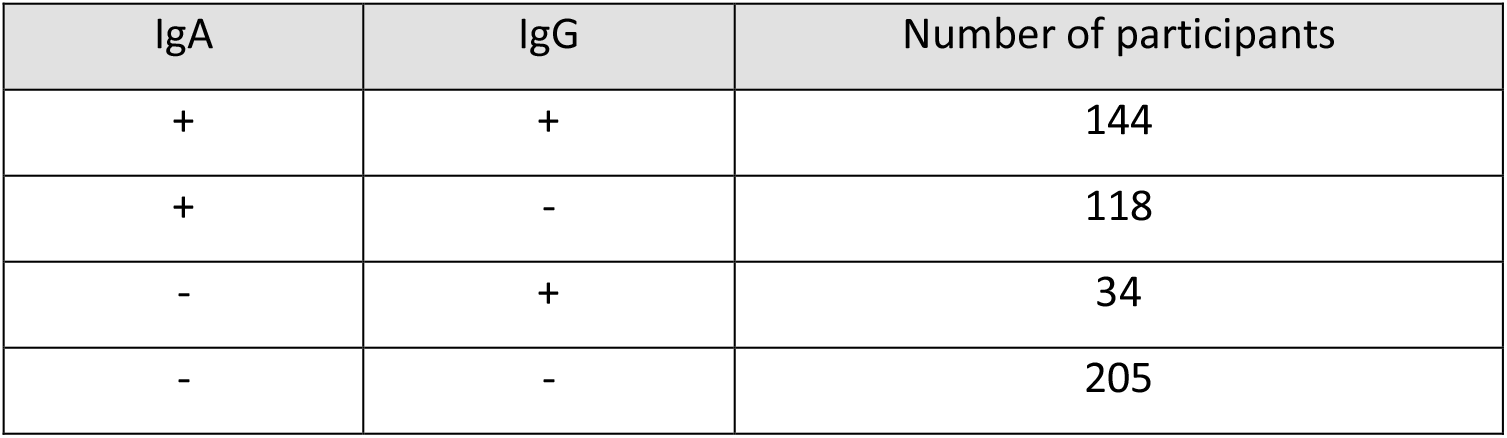
Results of qualitative testing for SARS-154 CoV-2-specific antibodies in healthy individuals (N=501), non-vaccinated and never diagnosed positively for SARS-CoV-2 infection

| IgA | IgG | Number of participants |
| --- | --- | --- |
| + | + | 144 |
| + | - | 118 |
| - | + | 34 |
| - | - | 205 |

These results in general are in line with the current epidemiological model developed by ICM University of Warsaw [7], where the percentage of recoveries in the population on April 1 was estimated at about 35% or 45%. Indeed, our study that was conducted later, during 15-22 May, revealed that half of the population may have been affected by SARS-CoV-2, as demonstrated by the unexpectedly high contribution of individuals positive for anti-SARS-CoV-2 IgA.

However, the virus-specific antibodies that confirm an individual’s contact or infection with the virus, do not necessary provide an effective protection from infections in the future. Anti-RBD IgG is considered the most significant fraction of virus-neutralizing antibodies, and it was found in 25.6% (128 out of 501) individuals, while 74.4% of the investigated population lacks anti-RBD IgG protection (Figure 1).

**Figure 1.**
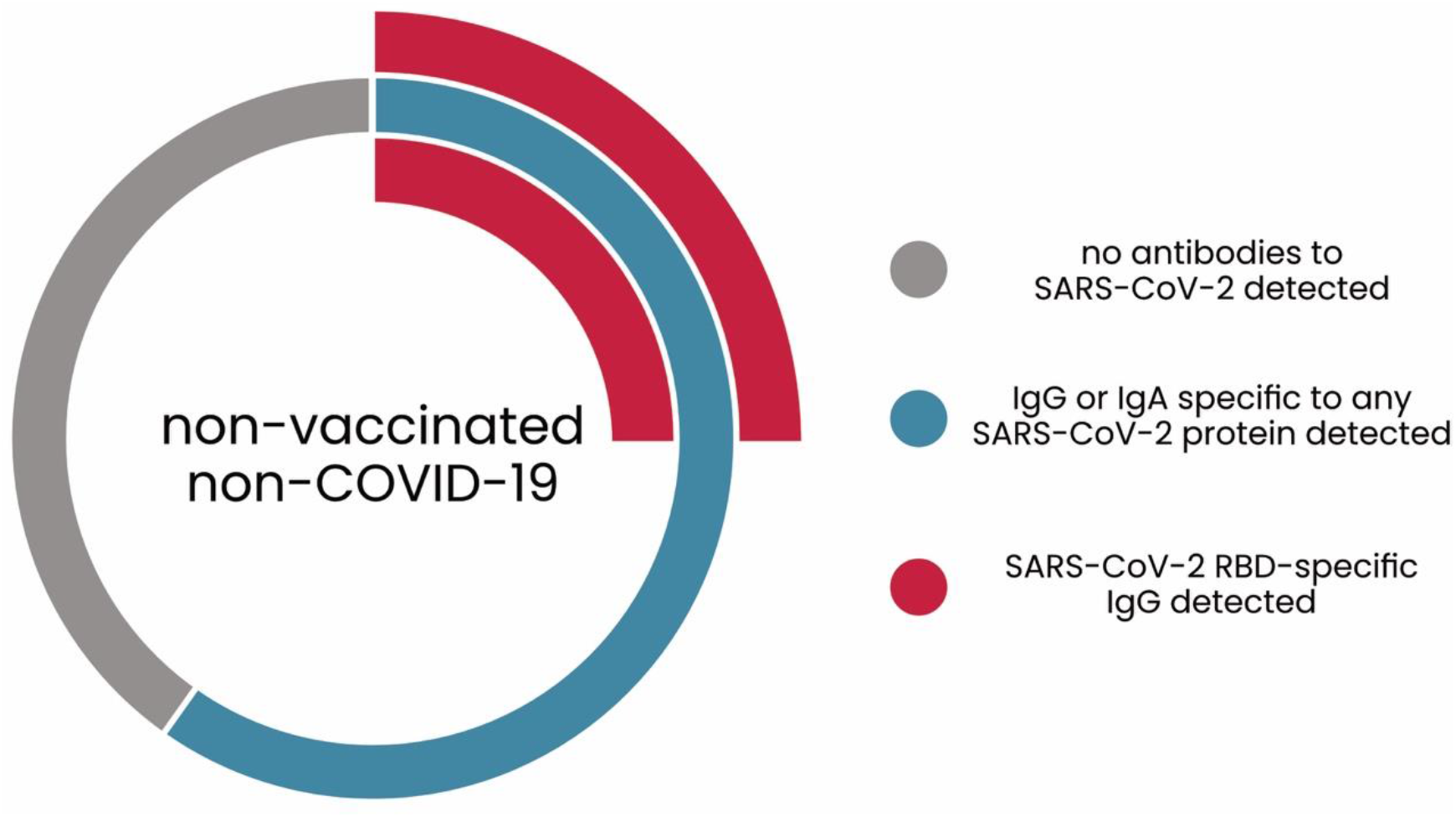
Fraction of the investigated population immune to SARS-CoV-2: patients positive for IgG and IgA.

We further compared quantitative levels of the virus-specific anti-RBD IgG in the fraction of healthy population identified as positive for anti-SARS-CoV-2 IgG, in COVID-19 patients hospitalized due to a severe course of the disease, and in individuals vaccinated against SARS-CoV-2. Median values were 525.12 U/ml, 1025.13 U/ml, and 954.93 U/ml, respectively, thus antibody levels were higher in the hospitalized patients or in the vaccinated individuals. These median values with quartiles and min-max values are presented in Figure 2.

**Figure 2.**
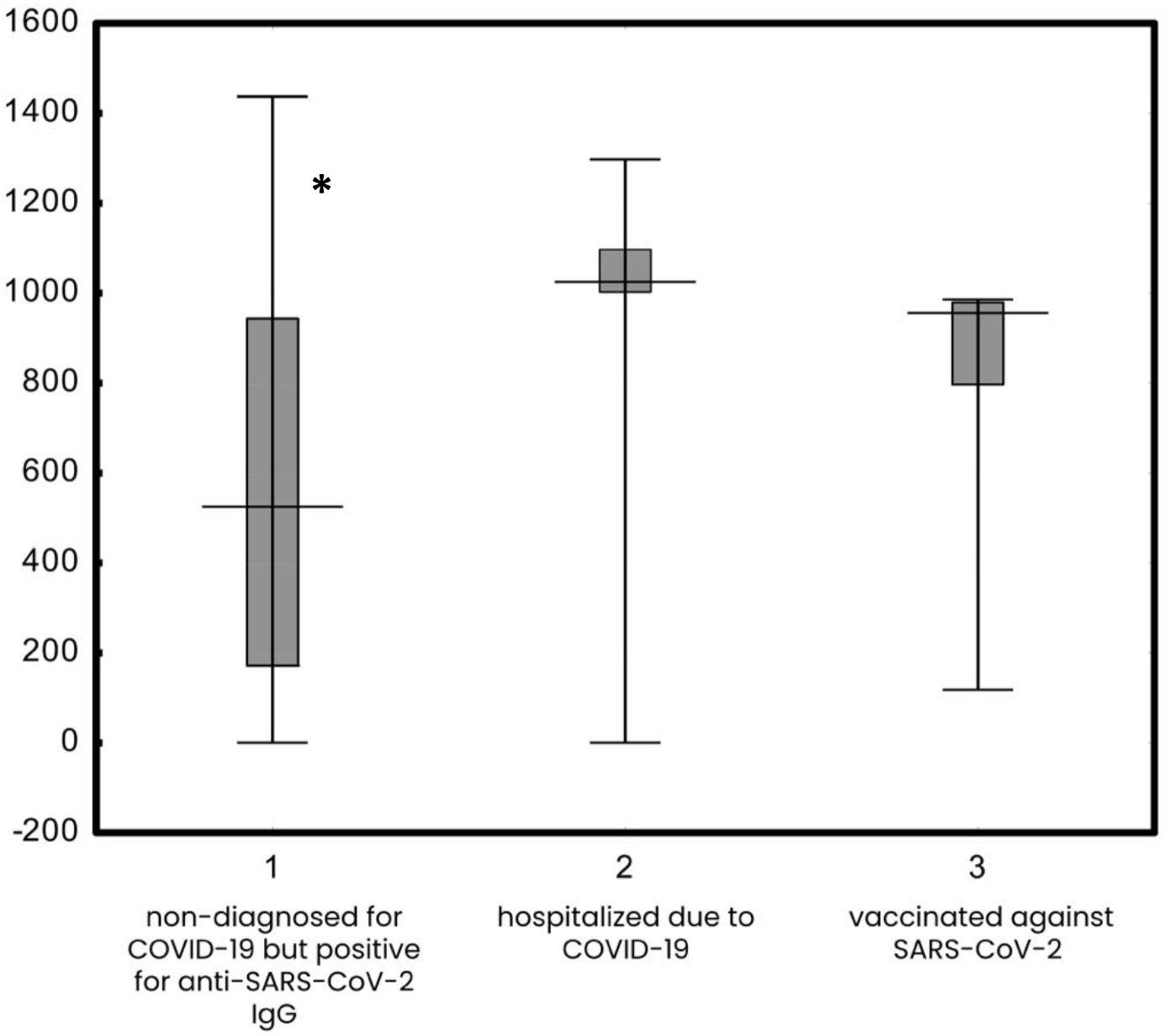
IgG specific to SARS-VoV-2 RBD in (1) non-diagnosed for COVID-19 but positive for anti-SARS-CoV-2 IgG, (2) in patients hospitalized due to COVID-19, (3) and in vaccinated against SARS-CoV-2. Median (vertical line), Quartiles (box), and Min-Max value (whiskers) of IgG antibody units per milliliter were presented; * p<0.05 when compared to hospitalized due to COVID-19.

To some extent, mucosal immunity can be provided by SARS-CoV-2-specific IgA. Within the group of SARS-CoV-2 IgA-positive patients, anti-RBD IgA was found in 136 individuals, which makes approximately 27.2% of the tested population.

This study has revealed a significant though still not sufficient fraction of the population of people who have developed natural immunological protection against COVID-19, apparently due to an asymptomatic, oligosymptomatic, or moderate (not-requiring hospitalization) course of the disease. Due to undiagnosed infections, they cannot be identified in the public health system as convalescents, being considered unaffected by SARS-CoV-2. This hidden fraction of immune individuals may really contribute to population immunity against COVID-19, improving the overall pandemic conditions and predictions [8]. On the other hand, this fraction seems to be insufficient for effective population immunity, since the majority of the investigated population still lacked anti-RBD IgG or IgA protection. Thus vaccination against COVID-19 is still of the utmost importance as the major tool for controlling the disease.

## Data Availability

We provide raw data applicable for any further analysis as the supplementary material

## Acknowledgments

This work was supported by the National Research and Development Center in Poland, grant no. SZPITALE-JEDNOIMIENNE/48/2020. The authors are deeply grateful to all participants who agreed to act as serum donors in this study. Thank you for your help, support, understanding, and for your desire to contribute to scientific solutions to combat the disease.

## Notes

### Competing Interest Statement

The authors have declared no competing interest.

### Funding Statement

This work was supported by the National Research and Development Center in Poland, grant no. SZPITALEJEDNOIMIENNE/48/2020

